# Clinical autism subscales have common genetic liabilities that are heritable, pleiotropic, and generalizable to the general population

**DOI:** 10.1101/2021.08.30.21262845

**Authors:** Taylor R. Thomas, Tanner Koomar, Lucas G. Casten, Ashton J. Tener, Ethan Bahl, Jacob J. Michaelson

## Abstract

The complexity of autism’s phenotypic spectra is well-known, yet most genetic research uses case-control status as the target trait. It is undetermined if autistic symptom domain severity underlying this heterogeneity is heritable and pleiotropic with other psychiatric and behavior traits in the same manner as autism case-control status. In N = 6,064 autistic children in the SPARK cohort, we investigated the common genetic properties of twelve subscales from three clinical autism instruments measuring autistic traits: the Social Communication Questionnaire (SCQ), the Repetitive Behavior Scale-Revised (RBS-R), and the Developmental Coordination Disorder Questionnaire (DCDQ). Educational attainment polygenic scores (PGS) were significantly negatively correlated with eleven subscales, while ADHD and major depression PGS were positively correlated with ten and eight of the autism subscales, respectively. Loneliness and neuroticism PGS were also positively correlated with many subscales. Significant PGS by sex interactions were found –– surprisingly, the autism case-control PGS was negatively correlated in females and had no strong correlation in males. SNP-heritability of the DCDQ subscales ranged from 0.04 to 0.08, RBS-R subscales ranged from 0.09 - 0.24, and SCQ subscales ranged from 0 to 0.12. GWAS in SPARK followed by estimation of polygenic scores (PGS) in the typically-developing ABCD cohort (N = 5,285), revealed significant associations of RBS-R subscale PGS with autism-related behavioral traits, with several subscale PGS more strongly correlated than the autism case-control PGS. Overall, our analyses suggest that the clinical autism subscale traits show variability in SNP-heritability, PGS associations, and significant PGS by sex interactions, underscoring the heterogeneity in autistic traits at a genetic level. Furthermore, of the three instruments investigated, the RBS-R shows the greatest evidence of genetic signal in both (1) autistic samples (greater heritability) and (2) general population samples (strongest PGS associations).

## 1 Introduction

Autism is a common and complex umbrella diagnosis affecting 1 in 59 children in the US [1], with comparable prevalence worldwide [2]. It has a strong genetic basis, with pedigree heritability estimated at 80% [3] and common genetic (SNP) heritability estimated at 0.13 [4]. *De novo* [5], inherited rare [6, 7], and inherited common genetic variation [4] can all contribute to autism liability, with *de novo*/rare and common genetic variants combining additively to increase risk [8, 9]. The DSM-5 defines autism by two core domains of symptomatology: the social domain, which includes social communication difficulties, and the non-social domain, which encompasses restricted and repetitive behaviors/interests [10]. However, other symptom domains are also characteristic and clinically meaningful in autism, including fine motor development [11], executive functioning [12], sleep [13], eating [14], and sensory issues [15]. Impaired fine motor skills are of particular interest due to being clinically detectable at 12 months, preceding deficits in the two core domains of autism, which emerge around two years of age [16]. Together, these multiple symptom domains imply that an accurate description of autism would be inherently multidimensional, rather than a simple binary diagnostic indicator. Because autism is strongly genetic, if these multiple symptom domains surpass the binary diagnosis in terms of their genetic signal, there may be broader implications for issues ranging from nosology to early identification [17] and individualized intervention [18].

However, most genetic research on autism has favored diagnostic status as the target trait. This is to be expected due to the relative ease of scaling a binary diagnostic phenotype to a large sample collected over many sites worldwide. Still, the traditional case-control approach in autism genetics is at odds with observations established by psychiatry and epidemiology in two major ways –– first by treating autism as a binary and homogeneous condition, and secondly by treating those without an autism diagnosis as true controls, despite the fact that autism is under-diagnosed (especially in females [19] and racial/ethnic minorities [20]) and many individuals without an autism diagnosis have autistic traits [21] [22]. While the case-control method has been successful in identifying a number of genes and biological processes associated with autism, variation in these genes has been found to represent broad neuropsychiatric risk with little specificity to autism [23]. As a result, it is possible that studies are losing statistical power to detect the genetic signal of core autistic traits by collapsing the clinical heterogeneity within cases and treating non-diagnosed individuals as true controls.

Across autistic individuals there is extreme variance in these core autistic traits and comorbid symptom domains, manifesting as heterogeneity in clinical presentation [24]. Clinical psychiatry has responded to this heterogeneity by developing numerous psychometric instruments to measure the social communication difficulties, restricted and repetitive behaviors/interests, and motor skills that are clinically relevant to autism. SPARK [25], a United States-based study of autism with over 270,000 individuals, collects a variety of information from participants, including background history, psychometric instruments, and other phenotypic information through online surveys. In this online setting, several clinically validated autism instruments, including the Social Communication Questionnaire (SCQ) [26], the Repetitive Behavior Scale-Revised (RBS-R) [27], and the Developmental Coordination Disorder Questionnaire (DCDQ) [28], have been completed by thousands of SPARK participants via self-report or parent-report. All three of these instruments and their subscales measure clinically relevant, quantitative traits that have important utility in the diagnosis of autism. Further investigation into the genetic characteristics of these clinical instruments could give insight into their best use cases as well as their potential limitations, potentially illuminating future autism diagnostic strategies and inherent biology.

Somewhat ironically, genetic investigation of quantitative autistic traits has gained the most traction in studies of the general population, yielding important insights. Most prominent are the recent genome-wide association studies (GWAS) of systemizing behavior, an indicator of restricted and repetitive behaviors/interests, (N = 51,564) [29], and empathy, an indicator of social communication abilities, (N = 46,861) [30]. They did not observe significant genetic correlations between systemizing and empathy, but did find greater systemizing and lower empathy were both significantly genetically correlated with autism (PGC) [29]. Another study in a general population (N = 1,981) derived five autistic factors, three of which were significantly associated with autism polygenic scores (PGS) [31]. These results suggest common genetic variation does play a significant role in quantitative autistic traits that case-control autism studies are inherently overlooking. However, the interchangeability of autistic traits from general population samples to autistic cohorts is unknown [32]. Results from genetic studies of autistic traits in autistic cohorts are limited by sample size and the availability of clinically validated instruments as phenotypes. One study in N = 2,509 autistic individuals found significant heritabilities and autism PGS associations with their autistic traits, but the phenotypes were derived factors based on items from the Autism Diagnostic Interview-Revised, not clinically established subscales measuring distinct symptomatology [33]. Therefore, the genetic etiology of autistic traits in autistic individuals is relatively indeterminate, especially with traits that have established clinical, psychological, and epidemiological usage in autism like the SCQ, RBS-R, and DCDQ subscales.

This analysis sought to investigate fundamental questions regarding the genetic etiology of dimensional autistic traits captured by twelve subscales from three of the primary autism clinical assessments. First, these subscales are designed to measure autistic traits that are meaningful at the clinical level, but are they measuring signal that is also meaningful at the genetic level in autistic individuals? Second, how heritable and genetically distinct are these quantitative traits, especially in an all-autistic cohort? Third, is the genetic signal discovered in this autistic cohort generalizable to related traits in a general population sample? Fourth, what are the pleiotropic relationships of these subscale traits with neuropsychiatric conditions, cognition, and dimensions of personality? Lastly, considering the observed sex differences that are pervasive in autism and other neuropsychiatric conditions, how do sex and PGS interact? We leveraged the SPARK autism cohort [25] to address these questions in N = 6,064 autistic children who all had the three DCDQ subscales, six RBS-R subscales, three SCQ subscales, and genetic data. An overview of our analyses is shown in Figure 1.

**Figure 1.**
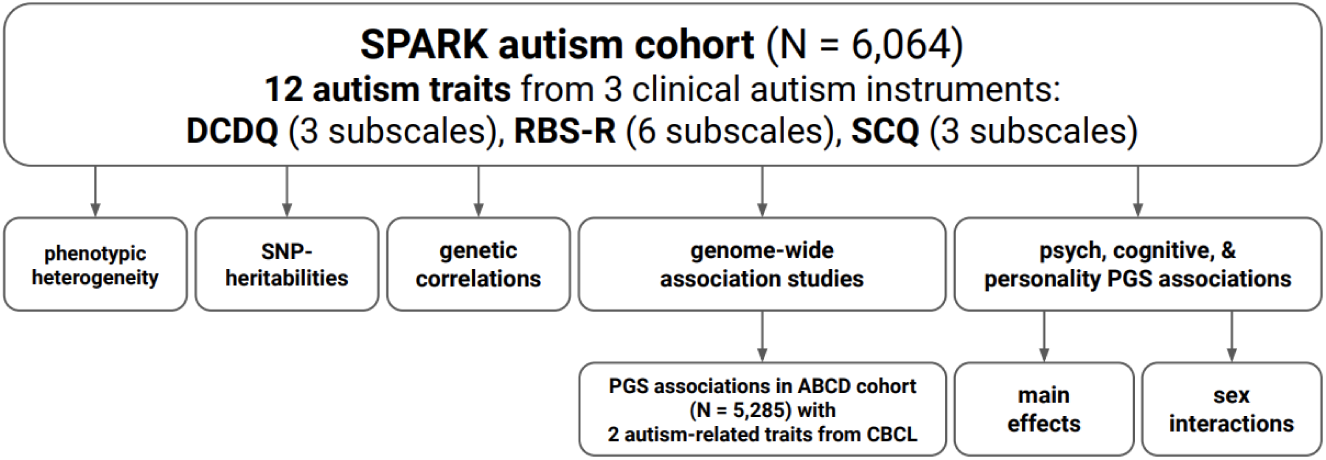
Study overview. Twelve quantitative, clinical autism subscale traits were measured in N = 6,064 autistic children in the SPARK cohort: three DCDQ subscales, six RBS-R subscales, and three SCQ subscales. These subscales were investigated both at the phenotype and genomic level, including SNP-heritabilities (*h*^2^), intra-cohort genetic correlations, genome-wide association studies (GWAS), and polygenic scores (PGS) associations with 14 psychiatric, cognitive, and personality traits. Additionally, the GWAS were used to calculate PGS in a general population childhood cohort, ABCD (N = 5,285). These PGS were then compared to the autism case-control PGS in association with two autism-related quantitative behavior traits from the CBCL.

## 2 Materials and methods

### 2.1 SPARK cohort description

SPARK [25] is a United States-based nationwide autism study. The parent or legal guardian of the child with autism provided informed consent and completed the psychometric instruments on behalf of their child. This study was approved by the Western IRB (IRB# 20151664). The SPARK Version 7 phenotype data release was used, and the Version 3 Freeze and Version 4 genotype data releases were used. We restricted the individuals in our analysis with the following criteria: has an autism diagnosis, is verbal (based on response to the first item in the Social Communication Questionnaire (SCQ)), was between the ages of 3 and 17 at the time the instrument was completed, passed the SPARK validity check for each instrument, and had full subscale scores for each of the twelve subscales. The cohort was further filtered to individuals in which genetic data was available that passed our quality control filtering, relatedness filtering, and were identified as European ancestry based on SNPs (genetic quality control, relatedness, and ancestry filtering described in further detail in the later section). After this filtering, we confirmed that the remaining individuals had unique family IDs. The final sample size of N = 6,064 was composed of N = 1,177 females and N = 4,887 males.

### 2.2 SPARK measures

#### 2.2.1 Primary phenotypes: clinical autism subscales

##### Developmental Coordination Disorder Questionnaire (DCDQ) [28]

a five point questionnaire of 15 items that is used clinically to screen for developmental coordination disorder. The DCDQ has three subscales: control during movement, fine motor/handwriting, and general coordination. These three subscales were calculated by SPARK and provided with the data release. The subscale scores within these scales were calculated as recommended by the scale publisher, including omitting the individual on a subscale due to missingness of item-level data within the subscale. In addition, the DCDQ was not completed if the participant was unable to move their hands. For the sake of consistent interpretation across the scales, the DCDQ scores were reversed so that higher scores indicated more coordination problems.

##### Restricted and Repetitive Behavior Scale-Revised (RBS-R) [27]

a four point questionnaire of 44 items that has been shown to be associated with several clinical features of autism [34]. The RBS-R has six subscales: compulsive behavior, self-injurious behavior, restricted behavior, ritualistic behavior, sameness behavior, and stereotyped behavior. These six subscales were calculated by SPARK and provided with the data release.

##### Social Communication Questionnaire (SCQ) [26]

a yes/no questionnaire of 40 items used as an autism screening tool in clinics [35]. We restricted the cohort to individuals with a ‘yes’ for the item 1 gate question: “uses phrases or sentences”. Item numbers 2, 9, 19, 20, 21, 22, 23, 24, 25, 26, 27, 28, 29, 30, 31, 32, 33, 34, 35, 36, 37, 38, 39, and 40 were reverse scored as recommended by the scale publisher. This was done to ensure that a 1 indicated increased severity for all the items used to calculate the subscales. The three SCQ subscales were calculated as outlined in [36]: communication, reciprocal social interaction, and stereotyped behavior.

In the sample of N = 6,064 individuals, the twelve subscale scores from these three assessments were first residualized for age in months using linear regression. Next, they were centered to have a mean of 0 and a standard deviation of 1. Finally, the scores underwent a rank-based normalization transformation [37] using the orderNorm() function from the bestNormalize R package [38]. These normalized scores were used as the phenotypes in all analyses.

#### 2.2.2 Secondary phenotypes

##### Child Behavior Checklist (CBCL) [39]

a three point questionnaire of 119 items used to assess behavior problems. The CBCL has eight syndrome subscales: anxious depressed, withdrawn depressed, somatic complaints, social problems, thought problems, attention problems, aggressive behavior, and rule-breaking behavior. From the final sample size (N = 6,064), a subset of N = 1,058 also had these subscale T-scores available, which were calculated by SPARK and provided with the data release. These CBCL subscales were first residualized for age in months using linear regression, then centered to have a mean of 0 and a standard deviation of 1, and then underwent a rank-based normalization transformation [37] using the orderNorm() function from the bestNormalize R package [38].

### 2.3 Phenotypic correlations and heterogeneity

The phenotypic correlations between each pair of subscales were calculated by the Pearson method, and the p-values were corrected using the Benjamini-Hochberg FDR [40] method. Cronbach’s alphas (a measurement of internal consistency or homogeneity) between each pair of subscales were calculated and then subtracted from 1 to represent the heterogeneity.

### 2.4 SPARK genotype quality control and imputation

Version 3 Freeze (2019) and Version 4 (2020) genotypes were first merged using PLINK [41]. The merged genotypes were then lifted from hg38 to hg19 using the LiftOver tool [42]. The genotypes included 43,209 individuals and 616,321 variants that were then quality controlled using the BIGwas quality control pipeline [43], which performed the genotype quality control, sample quality control, and identification of population stratification and sample filtering due to genetic ancestry. The BIGwas default parameters were used, except for skipping Hardy-Weinberg tests and keeping related samples due to the SPARK cohort being family-based and not a general population sample (we later removed related individuals with GCTA [44] within our SPARK sample of interest). The pre-QC annotation step removed 21 variants (N = 616,299 variants remaining). The SNP QC step removed 101,600 variants due to missingness at a threshold of 0.02 (N = 514,699 variants remaining). The sample QC step removed 1,114 individuals due to missingness, 67 individuals due to heterozygosity, and 176 due to duplicates (or monozygotic twins). The population stratification step projects the remaining individuals onto the principal components (PCs) from the combined HapMap and 1000 Genomes PCs and removed individuals who are not within median +/- five times the interquartile range for PC1 and PC2. This removed an additional 9,533 individuals (N = 32,422 individuals remaining). The quality controlled set of N = 514,699 variants and N = 32,422 individuals were then imputed to the TopMed [45] reference panel using the Michigan Imputation Server [46] with the phasing and quality control steps included and to output variants with imputation quality r2 > 0.3. After the genotype imputation, the variants were filtered to only the HapMap SNPs (N = 1,054,330 variants) with imputation quality r2 ≥ 0.8 using bcftools [47]. Next, they were lifted over from hg38 to hg19 using the VCF-liftover tool (https://github.com/hmgu-itg/VCF-liftover) and the alleles normalized to the hg19 reference genome. Finally, the files were converted to PLINK files with N = 1,018,200 final variants. This remaining set of quality controlled, imputed SNPs in the 32,422 individuals were filtered to the phenotype criteria as described in the above methods, and then removed for genetic relatedness with an identity-by-descent cutoff of 0.05 with SNPs at a minor allele frequency (MAF) ≥ 5% using GCTA [44], leaving the final sample size at N = 6,064.

### 2.5 SNP-based heritabilities and intra-cohort genetic correlations

GCTA [44] was used create a genetic relationship matrix for the N = 6,064 individuals (SNP MAF ≥ 5%). GCTA REML calculated the heritability for each subscale using sex as a covariate. GCTA Bivariate REML [48] calculated the genetic correlation between each pair of subscales also using sex as a covariate.

### 2.6 Genome-wide association studies and power

BOLT [49] version 2.3.5 was used to perform the genome wide association studies (GWAS) for each of the subscales using the BOLT-LMM option --lmm with sex as a covariate. The summary statistics for each GWAS were then filtered to only include SNPs in which the MAF ≥ 5%, leaving N = 941,385 SNPs. Lead SNPs (*p <* 5 × 10^−4^) were pruned using the PLINK [41] command --clump with default parameters and 1000 Genomes European as the LD reference. GWAS power was calculated using the genpwr.calc function from the genpwr R package [50] at several sample sizes. The average effect size (*β* estimate) and average MAF from the pruned SNPs for each GWAS were used to calculate power under an additive model.

### 2.7 ABCD cohort description

The ABCD cohort [51] is a typically-developing cohort was not recruited on the presence or absence of neuropsychiatric conditions. Release 2 data was used. We restricted the individuals in our analysis to those in which there were no missing data for the Child Behavior Checklist [39] Syndrome subscale T-scores. The cohort was then further filtered to individuals in which genetic data was available that passed our quality control filtering, relatedness filtering, and were identified as European ancestry based on SNPs (genetic quality control, relatedness, and ancestry filtering described in further detail in the later section). The final sample size of N = 5,285 was composed of N = 2,459 females and N = 2,826 males.

### 2.8 ABCD measures

#### Child Behavior Checklist (CBCL) [39]

a three point questionnaire of 119 items used to assess behavior problems. The CBCL has eight syndrome subscales: anxious depressed, withdrawn depressed, somatic complaints, social problems, thought problems, attention problems, aggressive behavior, and rule-breaking behavior. The two subscales items we used in our analyses were social problems and thought problems. The baseline intake year 1 scores were used. In the sample of N = 5,285 individuals, the CBCL subscales were first residualized for age in months using linear regression. Next, they were centered to have a mean of 0 and a standard deviation of 1.

Finally, the scores underwent a rank-based normalization transformation [37] using the orderNorm() function from the bestNormalize R package [38]. These normalized scores were used as the phenotypes in all analyses.

### 2.9 ABCD genotype quality control and imputation

The ABCD cohort was genotyped on the Affymetrix NIDA SmokeScreen Array and was processed through standard QC steps before release, including removing SNPs with low call rate and individuals with potential contamination problems or high missing data. The genotypes included 10,659 individuals and 517,724 variants that were then quality controlled using the BIGwas quality control pipeline [43], which performed the genotype quality control, sample quality control, and identification of population stratification and sample filtering due to genetic ancestry. The BIGwas default parameters were used, except for skipping Hardy-Weinberg tests in order to maintaining consistency with the SPARK BIGwas parameters (we also later removed related individuals with GCTA [44] within our ABCD sample of interest). The pre-QC annotation step removed 4,063 variants (N = 513,661 variants remaining). The SNP QC step removed 38,602 variants due to missingness at a threshold of 0.02 (N = 475,059 variants remaining). The sample QC step removed 434 individuals due to missingness, 12 individuals due to heterozygosity, and 336 due to duplicates (or monozygotic twins). The population stratification step projects the remaining individuals onto the principal components (PCs) from the combined HapMap and 1000 Genomes PCs and removed individuals who are not within median +/- five times the interquartile range for PC1 and PC2. This removed an additional 1,806 individuals (N = 8,115 individuals remaining). An additional N = 1,008 individuals were removed due to relatedness. The quality controlled set of N = 475,059 variants and N = 7,107 individuals were then imputed to the TopMed [45] reference panel using the Michigan Imputation Server [46] with the phasing and quality control steps included and to output variants with imputation quality r2 > 0.3. After the genotype imputation, the variants were filtered to only the HapMap SNPs (N = 1,054,330 variants) with imputation quality r2 ≥ 0.8 using bcftools [47]. Next, they were lifted over from hg38 to hg19 using the VCF-liftover tool (https://github.com/hmgu-itg/VCF-liftover) and the alleles normalized to the hg19 reference genome. Finally, the files were converted to PLINK files with N = 1,018,200 final variants. This remaining set of quality controlled, imputed SNPs in the 7,107 individuals were filtered to the phenotype criteria as described in the above methods, and then removed for genetic relatedness with an identity-by-descent cutoff of 0.05 with SNPs at a minor allele frequency (MAF) ≥ 5% using GCTA [44], leaving the final sample size at N = 5,285.

### 2.10 ABCD polygenic scores generation and statistical analyses

Polygenic scores (PGS) were calculated in the ABCD cohort using LDpred2 [52] and the bigsnpr tools [53] in R [54]. An external LD reference based on 362,320 European individuals of the UK Biobank (provided by the developers of LDpred2) was used to calculate the genetic correlation matrix, estimate heritability, and calculate the infinitesimal beta weights. The PGS were calculated from the filtered summary statistics for the subscale GWAS (MAF ≥ 5%) and the autism (PGC) [4] GWAS. The PGS were filtered to only include our sample of N = 5,285 and then the PGS were centered to have a mean of 0 and standard deviation of 1. PGS association with the CBCL phenotypes was assessed by a main effect linear model: lm(trait ∼ sex + PGS)). The sex is male coded as 1 and female coded as 0.

### 2.11 SPARK polygenic scores generation and statistical analyses

Polygenic scores (PGS) were calculated in the SPARK cohort using LDpred2 [52] and the bigsnpr tools [53] in R [54]. An external LD reference based on 362,320 European individuals of the UK Biobank (provided by the developers of LDpred2) was used to calculate the genetic correlation matrix, estimate heritability, and calculate the infinitesimal beta weights. Polygenic scores were calculated from the following genome-wide association studies performed by the Psychiatric Genomics Consortium: ADHD (2019) [55], anorexia nervosa (2019) [56], autism (2019) [4], bipolar disorder (2021) [57], major depression (2019) [58], schizophrenia (2020) [59], and Polygenic scores were calculated from genome-wide association studies performed by the Social Science Genetic Association Consortium for cognitive performance (2018) and educational attainment (2018) [60]. The public LDpred2 beta weights from the Polygenic Index Repository [61] were used to calculate polygenic scores for extraversion [62], neuroticism [63], openness [64], risky behavior [65], loneliness [66], and subjective well-being [67]. The PGS were then filtered to only include our sample of N = 6,064 and then the PGS were centered to have a mean of 0 and standard deviation of 1.

PGS associations with the subscales were analyzed by two complementary methods: linear modeling and Pearson correlations. The linear models tested for both main effects: lm(trait ∼ sex + PGS) and sex interactions: lm(trait ∼ sex + PGS + PGS : sex). The sex is male coded as 1 and female coded as 0. The PGS : sex is the PGS by sex interaction term. Correlations were tested within the entire sample (N = 6,064) and sex-stratified (N = 1,177 females and N = 4,887 males). For the grouped difference in means (autistic individual vs. parent) of the PGS, the cohort was filtered to autistic males in which their father also had genetic data available (N = 2,047 autistic males and N = 2,047 fathers) and autistic females in which their mother also had genetic data available (N = 638 autistic females and N = 638 mothers). The PGS of this subset were centered to have a mean of 0 and scaled to have a standard deviation of 1.

## 3 Results

### 3.1 Investigation of phenotypic correlations and heterogeneity

This study used the SPARK autism cohort [25], a United States-based nationwide cohort of autistic individuals and their parents and siblings. For our analyses of the autism subscales, we only included autistic children. The autistic traits used as the phenotypes were three subscales from the Developmental Coordination Disorder Questionnaire (DCDQ): coordination (general coordination), handwriting (fine motor handwriting), and movement (control during movement), six subscales from the Repetitive Behavior Scale-Revised (RBS-R): compulsive, injurious (self injurious), restricted, ritualistic, sameness, and stereotyped, and three subscales from the Social Communication Questionnaire (SCQ): communication, interaction (reciprocal social interaction), and stereotyped behavior. The demographic summary of the subset of SPARK used in this analyses and the raw subscales scores are shown in Table 1. The phenotypes used for all analyses in this study were these raw subscale scores that were residualized for age, centered to have a mean of 0 and a standard deviation of 1, and then normalized by rank-based normalization.

**Table 1.** Demographic and autism subscale summary of SPARK individuals in this study (N = 6,064). Ages in years.

| variable | % or mean (SD) |
| --- | --- |
| % male | 80% |
| cognitive imp. | 10% |
| dx age | 5 (2.7) |
| DCDQ age | 9.8 (3.2) |
| RBSR age | 9.7 (3.3) |
| SCQ age | 9.2 (3.3) |
| DCDQ coordination | 13.9 (4.3) |
| DCDQ handwriting | 9.9 (4.4) |
| DCDQ movement | 15.1 (5.9) |
| RBSR compulsive | 5.3 (4.2) |
| RBSR injurious | 3.4 (3.8) |
| RBSR restricted | 4.1 (2.9) |
| RBSR ritualistic | 6.2 (4.1) |
| RBSR sameness | 9.5 (6.4) |
| RBSR stereotyped | 5.3 (3.3) |
| SCQ communication | 6.5 (2.4) |
| SCQ interaction | 7.5 (3.7) |
| SCQ stereotyped | 6.4 (2) |

In order to assess the heterogeneity between subscales, we computed Pearson correlations and Cronbach’s alphas, with the results in Figure 2A and Table S1. The Pearson correlations of the twelve normalized subscales are shown in the red triangle in Figure 2A. All twelve of the subscales were significantly correlated with each other after FDR correction, although most correlations were modest. As expected, correlations were strongest between subscales within the same core domain. The highest Pearson correlation coefficient (*r*) was between RBS-R ritualistic and RBS-R sameness: *r* = 0.73. The weakest correlation was between DCDQ movement and RBS-R injurious: *r* = 0.13. Cronbach’s alpha (*a*) is a measure of internal consistency or homogeneity. While there is no definite cutoff of an *a* that indicates sufficient consistency/homogeneity, we considered any pair of subscales with an *a <* 0.7 to be heterogeneous [68]. Fifty-six of the 66 pairs of subscales were heterogeneous at this *a* cutoff, with the lowest *a* = 0.24 for DCDQ movement and RBS-R injurious. To represent the heterogeneity (instead of the homogeneity), the lower purple triangle in Figure 2A is showing 1 minus *a*. Figure 2B shows the pair of subscales with the lowest correlation and highest heterogeneity (left panel) and highest correlation/lowest heterogeneity (right panel).

**Figure 2.**
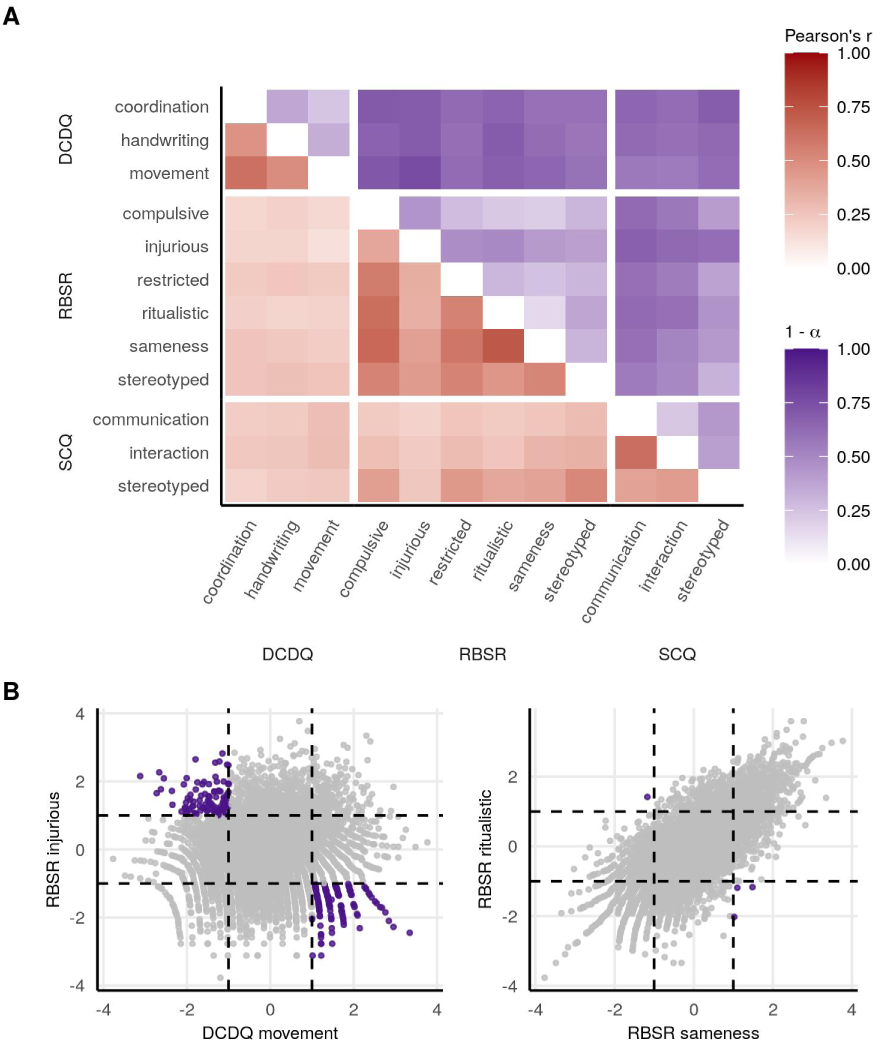
Phenotypic heterogeneity of the autism subscale traits. **A** The red triangle shows the Pearson correlation coefficient *r* between traits. All traits were significantly correlated with each other after FDR correction. The purple triangle shows the heterogeneity (1 minus Cronbach’s *a*). **B** The two subscales with the highest heterogeneity are on the left, and the two with the lowest heterogeneity are on the right. Individuals in purple are those who are ≥ 1 standard deviation above the mean in one subscale and *≤* 1 standard deviation below the mean in the other subscale.

Lastly, we correlated the twelve autistic trait subscales with the eight syndrome subscales from the Child Behavior Checklist (CBCL). The CBCL is a well-established tool that measures a range of behavior problems in children, and we wanted to determine which CBCL subscales were most correlated with the clinical autism subscales in anticipation of our later use of the CBCL subscales in our analysis the ABCD cohort. Therefore, in a subset of our SPARK sample (N = 1,058 of N = 6,064) in which we also had the CBCL subscales, we correlated the twelve autism subscales with the eight CBCL subscales. Table S2 has the correlations between the twelve autism subscales and the eight CBCL subscales. For each autism subscale, we found either social problems or thought problems to be the CBCL subscale most strongly correlated, in agreement with [69]. DCDQ coordination and DCDQ movement were most strongly correlated with CBCL social problems (both *r* = 0.17). The other ten subscales were most strongly correlated with CBCL thought problems, with the correlations ranging from *r* = 0.18 for both DCDQ handwriting and SCQ interaction to *r* = 0.41 for RBS-R injurious.

### 3.2 SNP-based heritabilities

Because our phenotypic analyses of the autism subscales identified the presence of heterogeneity and low correlations, we next wanted to identify the genetic factors underlying this heterogeneity. We first calculated additive SNP-based heritability (*h*^2^) for each subscale, which is the proportion of variance in the phenotype (subscale) that can be explained by the variance in the measured genetic data (the SNPs). We calculated the heritabilities using GCTA [44] and compared the heritabilities to the autism case-control GCTA heritability from the Psychiatric Genomics Consortium (PGC) [4]. Figure 3A shows the mean heritability estimate (*h*^2^) with the whiskers representing the 95% confidence intervals (CIs). Table S3 also includes the *h*^2^, standard error, 95% CIs around the estimate, and p-values. The overall heritability trend is that the RBS-R subscales have the highest heritabilities ranging from mean *h*^2^ of 0.09 to 0.24, the DCDQ subscales have moderate heritabilities from 0.04 to 0.08, and the SCQ subscales have the lowest heritabilities from 0 to 0.12. Despite large CIs due to the sample size, five of the six RBS-R traits were significantly heritable: RBS-R compulsive *h*^2^ = 0.21 (CI: 0.11 – 0.31), RBS-R injurious *h*^2^ = 0.20 (CI: 0.09 – 0.30), RBS-R restricted *h*^2^ = 0.24 (CI: 0.14 – 0.33), RBS-R sameness *h*^2^ = 0.21 (CI: 0.11 – 0.31), RBS-R stereotyped *h*^2^ = 0.19 (CI: 0.10 – 0.29). SCQ stereotyped was also significantly heritable: *h*^2^ = 0.12 (CI: 0.03 – 0.22). Notably, these five RBS-R traits has also have a mean *h*^2^ higher than the autism (PGC) heritability *h*^2^ = 0.13 (CI: 0.11 – 0.15) calculated by GCTA [4], although the 95% CIs around our *h*^2^ estimates are substantially larger due to much smaller sample size (the autism (PGC) sample size is N = 46,350 individuals), and the RBS-R CIs overlap with the autism (PGC) CIs.

**Figure 3.**
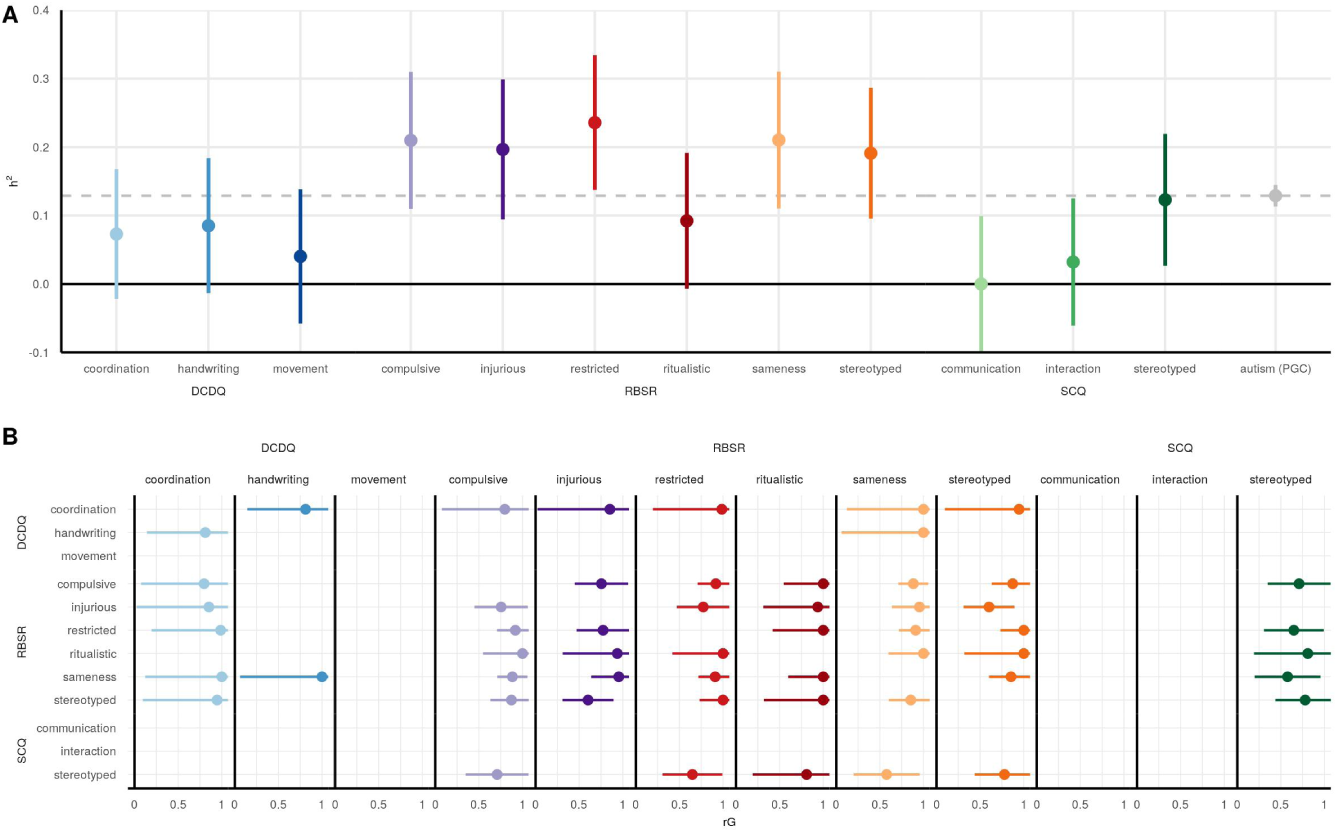
SNP heritabilities and genetic correlations. **A** SNP heritabilities (*h*^2^). The mean *h*^2^ estimate is plotted with the whiskers representing the 95% confidence intervals. For comparison, the autism case-control (PGC) heritability is also shown. **B** Intra-cohort genetic correlations (rG). The mean rG estimate is plotted with the whiskers representing the 95% confidence intervals. The rG value is plotted only if the correlation is nominally significant (unadjusted p-value *<* 0.05)

### 3.3 Genetic correlations

After observing differences in heritabilities across the twelve subscales, we next investigated the genetic correlations between the subscales. The genetic correlations (rG), also calculated with GCTA [48], are shown in Figure 3B with the mean rG and 95% CIs. Table S4 also includes the rG, standard error, 95% CIs around the estimate, and p-values. The rG standard errors are dependent on the heritability standard errors, and we did observe the more heritable traits having more significant genetic correlations. Therefore, only nominally significant (unadjusted p-value *<* 0.05) rG are reported in the figure. The overall trend was the strongest genetic correlations being within the same psychometric instrument/core domain. However, a few subscales in different domains were significantly genetically correlated. DCDQ coordination was positively genetically correlated with five of the RBS-R subscales, and DCDQ handwriting was positively genetically correlated with RBS-R sameness. SCQ stereotyped was positively genetically correlated with five RBS-R subscales.

### 3.4 Genome-wide association studies

Genome-wide association studies (GWAS) were conducted separately for the subscales using BOLT [49] and the SNPs were filtered to a minor allele frequency (MAF) ≥ 5%. The SCQ communication GWAS was unable to converge due to low heritability. The Manhattan plots are shown in Figure 4A. No SNPs reached genome-wide significance (*p <* 5 × 10^−8^). We therefore conducted a power analyses with the average effect size and MAF on a clumped set of lead SNPs (lead SNPs defined as *p <* 5 × 10^−4^) for each GWAS at five sample sizes: N = 6,064 (actual), N = 7,580 (actual times 1.25), N = 9,096 (actual times 1.5), N = 10,612 (actual times 1.75), and N = 12,128 (double). Table S5 has the number of lead SNPs used, the average *β* effect size, the average MAF, and the power at each N. Figure 4B is showing the power at each sample size for the least-powered GWAS from each of the three clinical instruments. At our sample size of N = 6,064, the power ranged from 16% to 26%. However, with double the sample size (N = 12,128), 80% power is reached or exceeded for all subscales, with power ranging from 81% to 91%.

**Figure 4.**
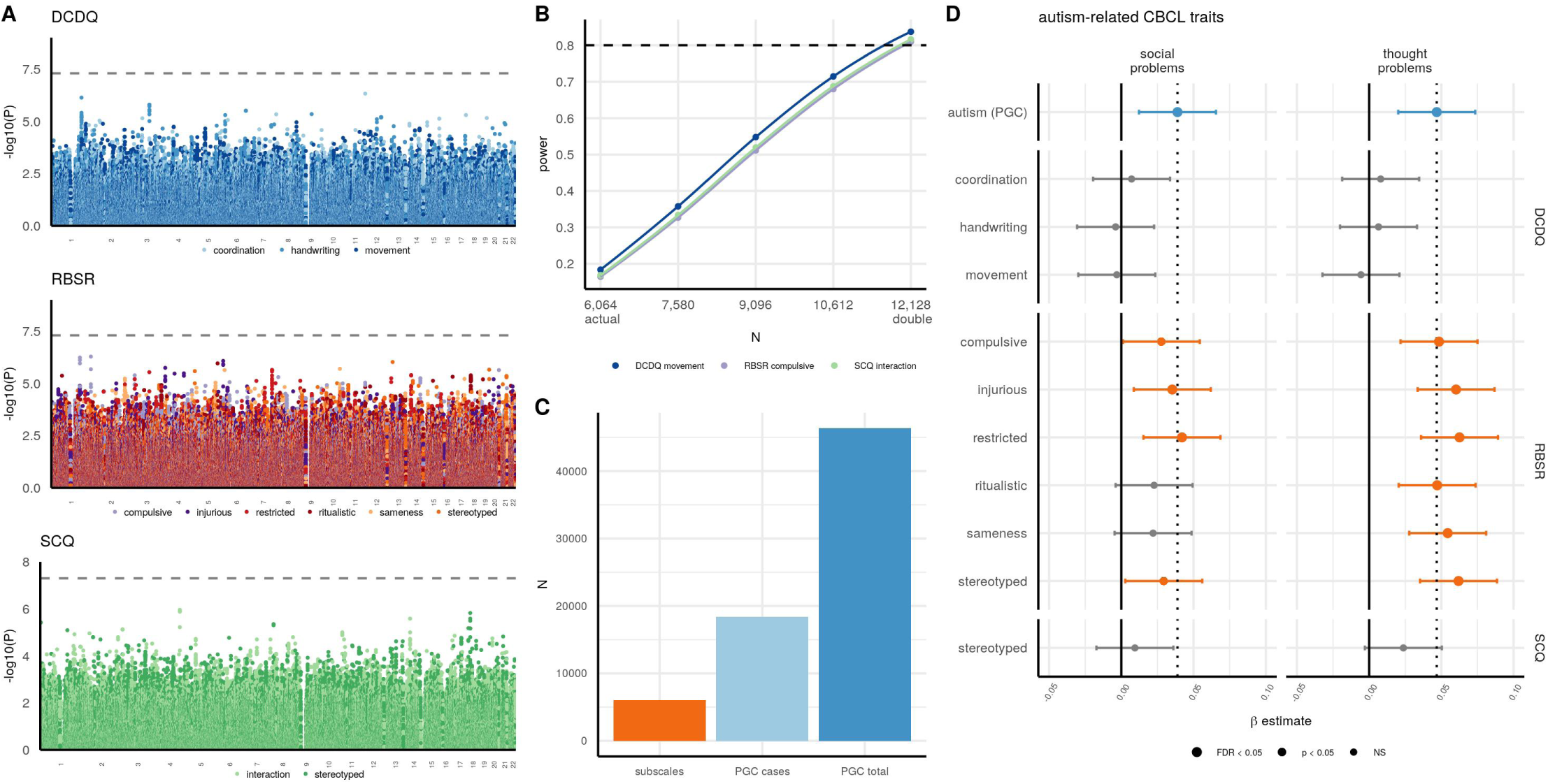
Subscale GWAS in SPARK and PGS estimations in ABCD associated with autism-related traits. **A** Manhattan plots for the GWAS of the subscales. The dashed line is the genome-wide significance line at *-log*_10_ (5 × 10^−8^). **B** Clumped lead SNPs (*p <* 5 × 10^−4^) for each GWAS were used to calculate power at sample sizes ranging from N = 6,064 (actual sample size) to N = 12,128 (double sample size). For each psychometric instrument/core domain, the subscale with the lowest power at N = 12,128 is shown. **C** Sample sizes of the subscale GWAS in comparison to the autism (PGC) cases and the autism (PGC) cases and controls. **D** PGS estimations from the subscale GWAS and the autism (PGC) GWAS were calculated in ABCD and associated with two CBCL measures shown to be elevated in autistic individuals–– social problems and thought problems [69]. The *β* estimate from the linear model is shown with the 95% confidence interval. The dashed line is the *β* estimate for the autism (PGC) PGS.

### 3.5 PGS estimations of subscale GWAS and their associations with behavior traits in ABCD

Despite our GWAS of the subscales not sufficiently powered for associations of individual loci, we calculated polygenic scores (PGS) in the ABCD cohort (N = 5,285) from each GWAS with LDpred2 [52] in order to test the generalizability of the genetic associations in a typically-developing sample. We tested the PGS associations with subscales from the Child Behavior Checklist (CBCL) [39], which measures behavior and emotional problems. The CBCL has eight syndrome subscales, but we focused our analyses on the two subscales we found to be most strongly correlated with each autism subscale, which were social problems and thought problems (see Results 3.1 and Table S2). Additionally, these two subscales have previously been shown to be three standard deviations higher in autistic children compared to undiagnosed children [69], providing further justification for selecting these two CBCL subscales as being most indicative of autistic traits. As a baseline comparison, we also calculated the PGS from the autism (PGC) GWAS. We expected the autism (PGC) GWAS to outperform the subscale GWAS because the autism (PGC) sample size of N = 46,350 is seven times greater than our sample (Figure 4C.)

The demographic summary of the ABCD cohort used for this analysis and the raw subscale values are shown in Table 2. The two CBCL measures were residualized for age using linear regression, centered to have a mean of 0 and a standard deviation of 1, and then normalized by a rank-based normalization. PGS performance was assessed using the main effects linear model lm(trait ∼ sex + PGS), with the *β* estimate from this model shown in Figure 4D along with the 95% CI around the *β*. The autism (PGC) PGS *β* estimate is shown with the dashed line. Table S6 has the *β* estimates, standard errors, 95% CIs, and p-values for the twelve subscales and autism (PGC) PGS associations with eight CBCL subscales.

**Table 2.**
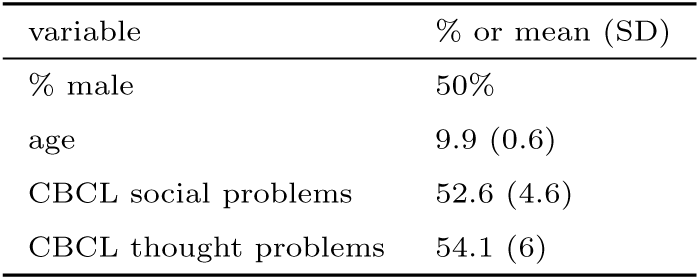
Autism-related CBCL behavior traits and demographic summary of ABCD individuals in this study (N = 5,285. Ages in years.

| variable | % or mean (SD) |
| --- | --- |
| % male | 50% |
| age | 9.9 (0.6) |
| CBCL social problems | 52.6 (4.6) |
| CBCL thought problems | 54.1 (6) |

The autism (PGC) PGS *β* estimate was significantly associated with CBCL social problems: *β* = 0.04 (CI: 0.01 – 0.07) and CBCL thought problems: *β* = 0.05 (CI: 0.02 – 0.07). Almost all of the RBS-R subscales were significantly associated with both CBCL social problems (compulsive *β* = 0.03, injurious *β* = 0.04, restricted *β* = 0.04, stereotyped *β* = 0.03) and CBCL thought problems (compulsive *β* = 0.05, injurious *β* = 0.06, restricted *β* = 0.06, ritualistic *β* = 0.05, sameness *β* = 0.05, stereotyped *β* = 0.06), with the RBS-R associations stronger for thought problems than social problems. PGS for SCQ interaction did not compute due to low LDSC heritability (LDpred2 requires LDSC heritability > 0), and no DCDQ subscales nor SCQ stereotyped were significantly associated with either CBCL subscale.

### 3.6 Polygenic scores main effects in SPARK

In order to assess pleiotropy of the autism subscales with neuropsychiatric conditions, cognitive traits, and behavior, we calculated polygenic scores (PGS) using LDpred2 [52] in the SPARK cohort from 14 publicly available GWAS: educational attainment (SSGAC) [60], cognitive performance (SSGAC) [60], ADHD (PGC) [55], anorexia (PGC) [56], autism (PGC) [4], bipolar disorder (PGC) [57], major depression disorder (PGC) [58], schizophrenia (PGC) [59], extraversion (PGI) [62], loneliness (PGI) [66], neuroticism (PGI) [63], openness (PGI) [64], risky behavior (PGI) [65], and subjective well-being (PGI) [67]. PGS main effect analyses were performed with two approaches: linear models of main effects lm(trait value ∼ sex + PGS) and Pearson correlations. The PGS main effects from the linear models are shown in Figure 5A with the *β* estimate and 95% confidence intervals (CIs) around the *β*. Table S7 has the *β* estimates, standard errors, 95% CIs, and p-values for PGS associations with the subscales. The Pearson correlation coefficients for the entire cohort and also sex-stratified correlations are in Table S8.

**Figure 5.**
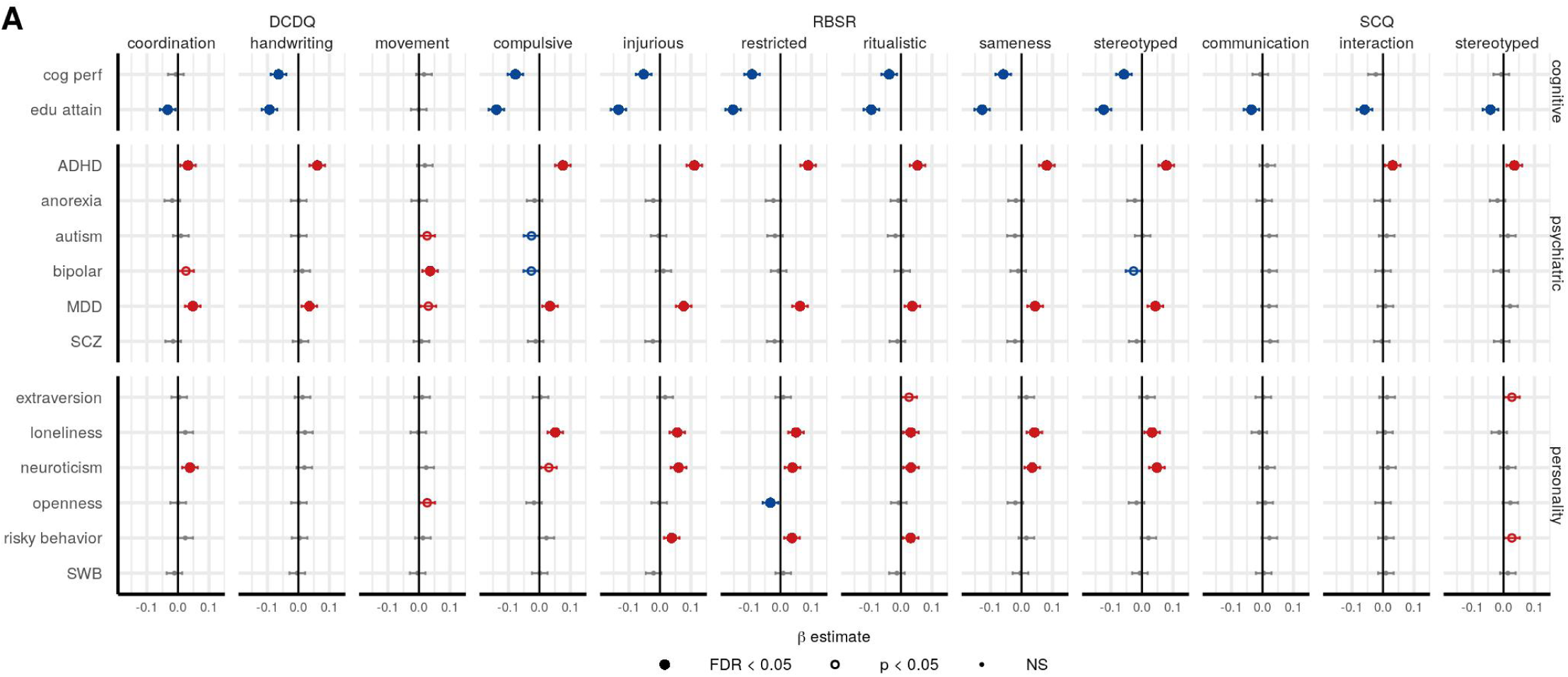
Main effects of polygenic scores in SPARK. **A** The PGS *β* estimate from the main effects modeling is plotted with the 95% confidence interval. Significant positive associations are in red, and significant negative associations are in blue, with a solid circle indicating significance after multiple testing correction (FDR *p <* 0.05) and an open circle indicating nominal significance only. PGS names are cog perf = cognitive performance, edu attain = educational attainment, MDD = major depression disorder, SCZ = schizophrenia, SWB = subjective well-being.

In general, educational attainment, cognitive performance, ADHD, and major depression had the most significant associations with the subscales. The strongest effects were seen in educational attainment PGS negatively associated with eleven subscales (FDR *p <* 0.05), including DCDQ coordination *β* = -0.03, DCDQ handwriting *β* = -0.09, RBS-R compulsive *β* = -0.14, RBS-R injurious *β* = -0.13, RBS-R restricted *β* = -0.15, RBS-R ritualistic *β* = -0.10, RBS-R sameness *β* = -0.13, RBS-R stereotyped *β* = -0.12, SCQ communication *β* = -0.04, SCQ interaction *β* = -0.06, SCQ stereotyped *β* = -0.04. This effect was also seen in most of the cognitive performance PGS as well, meaning higher PGS for cognitive performance and educational attainment are associated with reduced autistic traits. Surprisingly, the autism (PGC) PGS only had only a slight positive association with DCDQ movement *β* = 0.03 and a slight negative association with RBS-R compulsive *β* = -0.03, neither of which was significant after FDR. In contrast, ADHD PGS was positively associated with ten subscales (FDR *p <* 0.05): DCDQ coordination *β* = 0.03, DCDQ handwriting *β* = 0.06, RBS-R compulsive *β* = 0.08, RBS-R injurious *β* = 0.11, RBS-R restricted *β* = 0.09, RBS-R ritualistic *β* = 0.05, RBS-R sameness *β* = 0.08, RBS-R stereotyped *β* = 0.08, SCQ interaction *β* = 0.03, and SCQ stereotyped *β* = 0.04. Likewise, the major depression PGS was positively associated with all three DCDQ subscales and all six RBS-R subscales (FDR *p <* 0.05): DCDQ coordination *β* = 0.05, DCDQ handwriting *β* = 0.04, DCDQ movement *β* = 0.03, RBS-R compulsive *β* = 0.03, RBS-R injurious *β* = 0.08, RBS-R restricted *β* = 0.06, RBS-R ritualistic *β* = 0.04, RBS-R sameness *β* = 0.04, and RBS-R stereotyped *β* = 0.04. The bipolar disorder PGS was positively associated (FDR *p <* 0.05) with DCDQ movement *β* = 0.04.

We also found significant personality PGS associations with the subscales. The neuroticism PGS was positively associated with seven subscales (FDR *p <* 0.05): DCDQ coordination *β* = 0.04, RBS-R compulsive *β* = 0.03, RBS-R injurious *β* = 0.06, RBS-R restricted *β* = 0.04, RBS-R ritualistic *β* = 0.03, RBS-R sameness *β* = 0.04, and RBS-R-stereotyped *β* = 0.05. The loneliness PGS was positively associated with all six RBS-R subscales (FDR *p <* 0.05): RBS-R compulsive *β* = 0.05, RBS-R injurious *β* = 0.06, RBS-R restricted *β* = 0.05, RBS-R ritualistic *β* = 0.03, RBS-R sameness *β* = 0.04, RBS-R stereotyped *β* = 0.03. The risky behavior PGS was positively correlated with three RBS-R subscales (FDR *p <* 0.05): RBS-R injurious *β* = 0.04, RBSR-R restricted *β* = 0.04, RBS-R ritualistic *β* = 0.03.

### 3.7 Interaction of sex with polygenic scores in SPARK

In order to analyze sex interactions of the autism subscale traits with neuropsychiatric, cognitive, and personality PGS, we first formally tested for sex interaction effects by linear models with a PGS by sex interaction term lm(trait ∼ sex + PGS + PGS : sex). We observed 11 nominally significant PGS by sex interaction effects. Figure 6A is showing the *β* estimate from the PGS by sex interaction term. Table S9 has the *β* estimates, standard errors, 95% CIs, and p-values for PGS by sex interaction terms. While this interaction modeling allows identification of significant PGS by sex interaction effects, it does not show how the interaction is manifested within each sex. For example, a positive *β* estimate for the PGS by sex interaction term (note that male was coded as 1 and female was coded as 0) can be indicative of one of three possible mechanisms:

1. The PGS has a positive effect in males and no effect in females.
2. The PGS has a positive effect in males and negative effect in females.
3. The PGS has no effect in males and a negative effect in females.

**Figure 6.**
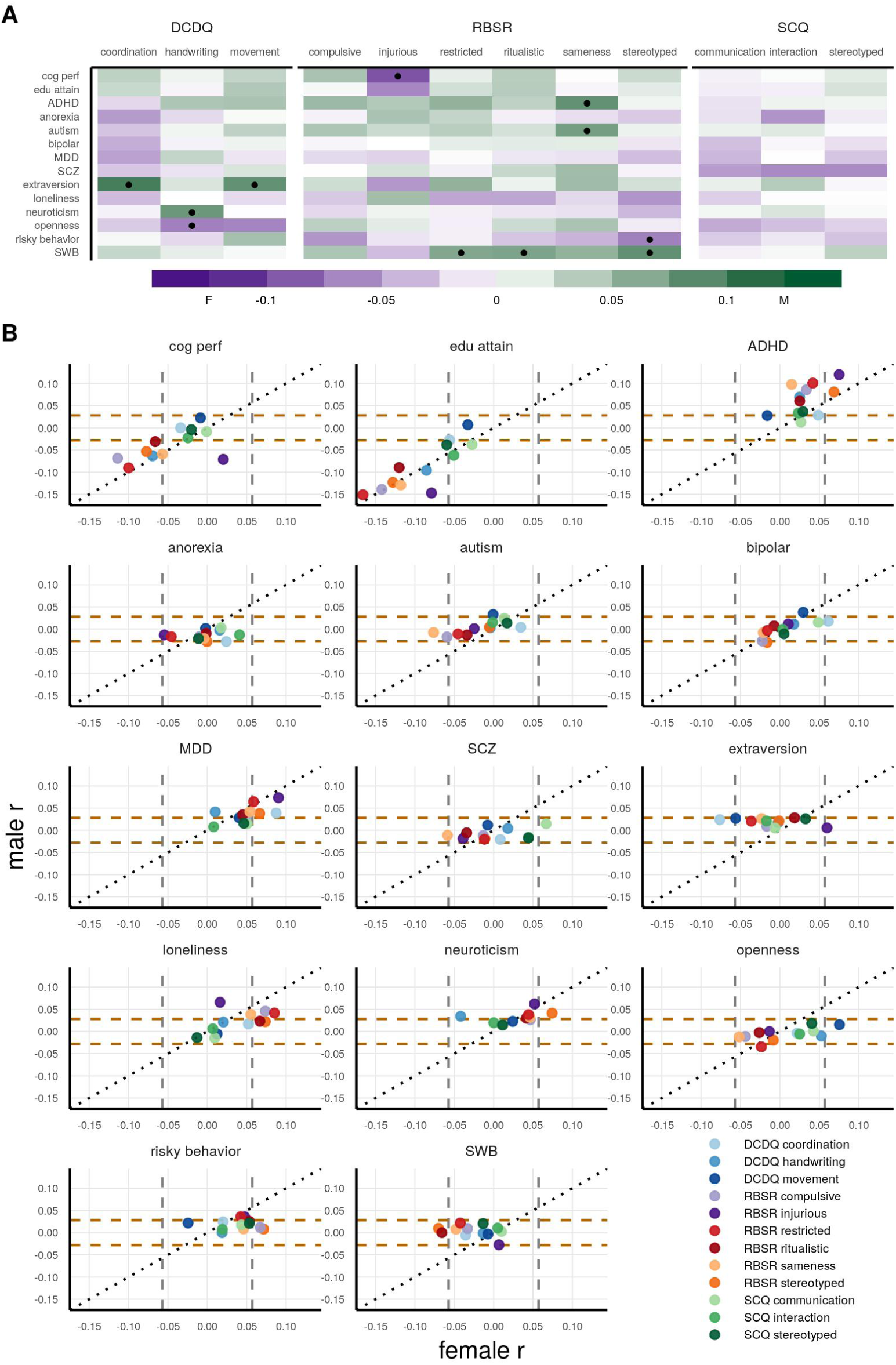
PGS sex interactions and sex-stratified correlations with the autism subscales in SPARK. **A** The PGS *β* estimate from the sex interaction modeling is shown with the fill color, with one dot indicating the PGS by sex interaction term was nominally significant. A positive *β* (green) indicates higher PGS by sex interaction for males, whereas a negative *β* (purple) indicates higher PGS by sex interaction for females. **B** Pearson *r* coefficient correlations for the female correlation (x-axis) vs. male correlation (y-axis) are plotted. The gray vertical lines indicate the bounds of nominal significance for females, and the brown horizontal lines indicate the bounds for males. N = 1,177 females and N = 4,887 males. PGS names are cog perf = cognitive performance, edu attain = educational attainment, MDD = major depression disorder, SCZ = schizophrenia, SWB = subjective well-being.

Likewise, the three possible interpretations of a negative *β* estimate (female direction) for the interaction term are:

1. The PGS has a positive effect in females and no effect in males.
2. The PGS has positive effect in females and negative effect in males.
3. The PGS has no effect in females and a negative effect in males.

Therefore, to further interrogate the sex interactions, we performed sex-stratified Pearson correlations (Figure 6B and Table S8). The significance of these correlations are important to understand in the context that there are four times as many males than females in the cohort (N = 1,177 females and N = 4,887 males). Figure 6B shows the Pearson *r* for the females vs. the *r* for males, with the dashed lines indicating the bounds of nominal significance (*p <* 0.05) for each sex-stratified correlation (*r* = +/- 0.06 for females and *r* = +/- 0.03 for males). The null hypothesis is the PGS effect to be the same for males and females, which is represented by the black diagonal line.

Overall, we observed several PGS having sex interaction effects and sex-differential correlations. Cognitive performance and educational attainment PGS did not have sex interaction effects, except for cognitive performance with RBS-R injurious (*β* = -0.09), with no correlation in females (*r* = 0.02) and a negative correlation in males (*r* = -0.07). In general, the ADHD PGS had stronger positive correlations in males than in females, e.g. RBS-R sameness had no correlation in females (*r* = 0.02) and a strong correlation in males (*r* = 0.10), interaction *β* = 0.08. Similar trends were observed for RBS-R compulsive: female (*r* = 0.03) and male (*r* = 0.09) and RBS-R restricted: female (*r* = 0.04) and male (*r* = 0.10). Surprisingly, we observed a negative correlation in females and no correlation in males with several RBS-R subscales and autism (PGC) PGS, meaning increasing PGS for autism had approximately no association with RBS-R traits in males and is actually associated with reduced RBS-R traits in females. The strongest sex interaction effect was for RBS-R sameness (interaction *β* = 0.07), with a negative correlation in females (*r* = -0.08) and no correlation in males (*r* = -0.01). Similar trends were observed for RBS-R compulsive (interaction *β* = 0.04): female (*r* = -0.06) and male (*r* = -0.02), and also for RBS-R restricted (interaction *β* = 0.04): female (*r* = -0.04) and male (*r* = -0.01). While no significant sex interactions for bipolar and major depression PGS, we did observe sex-differential correlations. Bipolar PGS was positively correlated with DCDQ coordination in females (*r* = 0.06) but the correlation was much less in males (*r* = 0.02). The same trend was observed for major depression PGS more strongly positively correlated in females for DCDQ coordination (female *r* = 0.09 and male *r* = 0.04) and RBS-R stereotyped (female *r* = 0.07 and male *r* = 0.04). For the schizophrenia PGS, we observed a stronger positive correlation in females (*r* = 0.07) than in males (*r* = 0.01) with SCQ communication.

The extraversion PGS had significant sex interactions with DCDQ coordination (*β* = 0.10) and DCDQ movement (*β* = 0.08), with negative correlations in females and a slight positive correlation in males. Interestingly, for RBS-R injurious and extraversion PGS, higher PGS was positively correlated in females (*r* = 0.06), but had no correlation in males (*r* = 0.01). We did not observe any significant loneliness PGS by sex interactions, although some showed sex-differential correlations. For example, RBS-R stereotyped was positively correlated in females (*r* = 0.07) but no correlation in males (*r* = 0.02). For the neuroticism PGC, there was a significant sex interaction with DCDQ handwriting (*β* = 0.08), with a negative trend in females (*r* = -0.04) and a positive correlation in males (*r* = 0.03). The risky behavior PGS and subjective well-being PGS had stronger correlations in females than males. Risky behavior PGS and RBS-R compulsive were positively correlated in females (*r* = 0.07) but not in males (*r* = 0.01), and likewise for RBS-R stereotyped (female *r* = 0.07 and male *r* = 0.01). The subjective well-being PGS were negatively correlated in females but not in males for RBS-R stereotyped (female *r* = -0.07 and male *r* = 0.01) and RBS-R ritualistic (female *r* = -0.07 and male *r* = 0).

Lastly, to further interrogate the complex relationship between autism (PGC) PGS and sex, we analyzed the grouped means of the autistic males for whom their fathers also had genetic data available (N = 2,047 autistic males and N = 2,047 fathers) and autistic females for whom their mothers also had genetic data available (N = 638 autistic females and N = 638 mothers). We expected autism (PGC) to perform well in predicting case-control status in SPARK given that previous work performed an autism case-control GWAS in SPARK and found the genetic correlation with autism case-control (PGC) to be high [70]. Indeed, we found that the autistic females had significantly higher autism (PGC) PGS than their mothers, and likewise the autistic males had significantly higher PGS than their fathers (Table 3), with the mean difference between between autistic females vs. mothers and autistic males vs. fathers being similar at 0.15 for females and 0.17 for males. However, the autistic females overall have higher PGS, with the autistic female mean at 0.13 and the autistic male mean at 0.07. Likewise, the mother mean is -0.02 and the father mean is -0.10.

**Table 3.** Difference in grouped mean PGS between the autistic individuals and their sex-matched parent. The cohort was filtered to autistic males in which their father also had genetic data available (N = 2,047 autistic males and N = 2,047 fathers) and autistic females in which their mother also had genetic data available (N = 638 autistic females and N = 638 mothers).

| PGS | sex | role | mean | mean dif |
| --- | --- | --- | --- | --- |
| ADHD | F | autistic | 0.05 | 0.03 |
|  |  | parent | 0.02 |  |
|  | M | autistic | 0.03 | 0.08 |
|  |  | parent | -0.05 |  |
| autism | F | autistic | 0.13 | 0.15 |
|  |  | parent | -0.02 |  |
|  | M | autistic | 0.07 | 0.17 |
|  |  | parent | -0.10 |  |
| edu attain | F | autistic | 0.01 | 0.13 |
|  |  | parent | -0.12 |  |
|  | M | autistic | 0.04 | 0.04 |
|  |  | parent | 0.00 |  |
| MDD | F | autistic | 0.08 | -0.01 |
|  |  | parent | 0.09 |  |
|  | M | autistic | 0.03 | 0.1 |
|  |  | parent | -0.08 |  |

## 4 Discussion

Our central goal for this work was to begin to reconcile autism’s strong genetic basis with its extensive phenotypic heterogeneity by examining the genetic characteristics of twelve subscales from widely-used clinical instruments (SCQ, RBS-R, and DCDQ). We employed well-established approaches such as genome-wide association studies, SNP-heritability estimations, and polygenic score associations. The findings from our primary analysis of N = 6,064 autistic children from SPARK and our generalization analysis of N = 5,285 children from the community-based cohort ABCD, which are detailed below, show pronounced differences in genetic signal underlying the three assessments we investigated. These findings also underscore the limitations in signal strength (heritability) and in generalizability of genetic associations that are based on a binary diagnosis. Taken together, our results show that the future of autism genetics demands a focus on multidimensional, quantitative phenotypes.

Our phenotypic analyses demonstrate the quantitative and heterogeneous nature of core autistic traits in an autism-only cohort. Some traits were only moderately correlated with each other, especially subscales probing different core domains (Figure 2A). In addition, at a Cronbach’s alpha cutoff of *a* = 0.7, 56 out 66 pairs of traits were heterogeneous. Autism is often described as a heterogeneous condition [71], and these findings serve to quantify that heterogeneity. More detailed phenotypic characterization and new methods for integrating these granular phenotypes [72] into robust, interpretable traits are critical goals as the field strives to develop a more actionable, mechanistic understanding of autism that is based on genetics.

Clinicians administering screening tools like the SCQ, RBS-R, and DCDQ might well wonder to what extent these instruments are detecting behaviors and traits that have bases in biology. Our analysis of SNP-heritability of each of these scales sheds light on such questions: a screen that is sensitive to relevant genetic factors should have higher heritability than a screen that is more environmentally influenced or more sensitive to state vs. trait distinctions. We found that SNP-heritability varied widely across the twelve subscales from 0 for SCQ communication to 0.24 for RBS-R restricted (Figure 3A). Five of the RBS-R subscales were significantly heritable and more heritable than autism (PGC) case-control at 0.13, although the confidence intervals around our estimates are much greater and overlap with the autism case-control confidence intervals (due to our sample size of N = 6,064 being seven times smaller than the autism (PGC) sample size of N = 46,350). SCQ stereotyped was also significantly heritable at 0.13, while the DCDQ subscales ranged from 0.04 to 0.08 and did not reach significance. Overall, these results suggest that the RBS-R is more sensitive to relevant genetic factors than either the SCQ or DCDQ.

Given the recent progress in parsing the genetics of latent autistic traits in large undiagnosed samples [29] [30], we were motivated to see if we could uncover evidence running in the other direction: are PGS based on our analysis of the twelve subscales in an autism-only cohort positively correlated with autism-related phenotypes in a general population sample of children? We calculated PGS in ABCD (the general population sample) using our subscale GWAS, and correlated these estimates with two quantitative behavioral traits from the CBCL that have previously been demonstrated to have a strong association with autism: social problems and thought problems [69]. Almost all the RBS-R subscale PGS were significantly associated with both social problems and thought problems, as was the autism (PGC) case-control PGS (Figure 4D), despite our GWAS sample size being much smaller and underpowered for locus discovery. This is in line with a previous study that also found an autism case-control PGS was significantly associated with social communication difficulties in a general population sample of N = 5,628 [73].

In association analyses with the autism case-control PGS to the subscales in SPARK, we hypothesized that the autism case-control PGS would have a positive association with the subscales. Surprisingly, we observed no significant associations after FDR (Figure 5). This may be due to the autism (PGC) GWAS collapsing the heterogeneity in the autism cases, which has the effect of pooling the autism risk alleles broadly. This is in agreement with previous research, which observed the autism case-control PGS to not lack significant association with IQ [9], the Autism Diagnostic Observation Schedule, Autism Diagnostic Interview-Revised, RBS-R, or Social Responsiveness Scale total scores in autistic individuals [74]. Interestingly, we found that when stratified by sex, the autism case-control PGS exhibits different associations with severity in males and females (Figure 6B), with the PGS having a correlation of roughly zero in males and being anti-correlated with symptom severity in females. This unexpected finding warrants some discussion of potential explanations and, in particular, sex-specific explanations. It has been previously established that affected children (mostly sons) inherit more genetic autism risk from their mothers [75]. Because the autism PGC cohort is heavily male-biased, the subset of risk alleles identified using this cohort can be seen as representing disproportionate “mother-to-son” alleles or male-specific risk alleles. Alternatively, these may be alleles that are female-benign, meaning they do not contribute to female risk; or female-protective, diminishing risk in females. The presence of these male-specific or female-benign/protective alleles can provide one explanation for why the autism PGS is correlated with lower symptom severity in females. On the other hand, autistic females carry more rare variant burden than autistic males [76]. It has been established that PGS and rare variants interact additively in autism risk [8, 9]. It is possible that the most severely affected autistic females in our cohort carry more rare variants that impact their severity, as recent analyses by [77] suggest. These two explanations are, of course, not mutually exclusive. Together, they present a strong case that it is crucial to consider sex in PGS applications, especially in sex-imbalanced cohorts where a sex-biased trait is studied [78].

Finally, we found that beyond autism, other neuropsychiatric, cognitive, and personality PGS were significantly associated with autistic traits in autistic individuals (Figure 5 and Figure 6B). The strongest PGS associations we found were educational attainment and cognitive performance PGS being negatively correlated with several subscales, meaning higher polygenic propensity for cognitive abilities in autistic individuals is associated with reduced autistic traits. Interestingly, this contrasts with numerous reports of a positive genetic correlation between educational attainment and autism case-control (PGC) [4, 79, 80], but it is in agreement with previous work showing higher PGS for educational attainment was positively associated with greater IQ in autistic individuals [9]. The ADHD and major depression PGS were both positively correlated with several subscales, which is in line with previous work showing ADHD and major depression to be positively genetically correlated with autism [4]. Additionally, factor analysis of genetic correlations across the major psychiatric disorders identified major depression to have a strong loading with the neurodevelopmental factor (ADHD and autism) [81], and when including substance use disorders, ADHD, autism, and major depression load onto the same factor [82]. This relationship of autism with ADHD and major depression is also shown at the epidemiological level, with ADHD and major depression being the first and third most prevalent comorbidities among autistic individuals, respectively [83]. Therefore, it is possible that the ADHD and major depression PGS in our analyses are indexing the autism phenotypic spectrum and/or subgroup comorbidities. Beyond neuropsychiatric and cognitive PGS, we also wanted to assess whether personality traits like extraversion, neuroticism, openness, and risky behavior PGS were also associated with the autism subscales. There is extensive research on the Big Five personality measures and their relationship to autism [84] and subtyping autistic individuals [85], but it is unknown if the genetic signals of personality can also subtype autistic individuals. Indeed, several personality PGS were associated with many of autism subscales (Figure 5). Among them, most prominently, several RBS-R subscales were positively associated with neuroticism, loneliness, and risky behavior PGS. Again sex was an important variable, with significant PGS by sex interactions observed for seven of the personality PGS (Figure 6). Overall, these analyses suggest that optimizing PGS as predictors, e.g. for eventual use in personalizing care, will require the utilization of several neuropsychiatric, cognitive, and personality PGS, as well as consideration of interactions with sex.

Our analyses have several limitations. First, our moderate sample size of N = 6,064 autistic children in SPARK is low for common genetic variant analyses, specifically for estimations of SNP-heritability, intra-cohort genetic correlations, and genome-wide association studies. This is clear from the wide confidence intervals for our heritability and genetic correlations (Figure 3), as well as the power analyses for our GWAS ranging from 19% to 22% (Figure 4B). This sample size limitation is especially important to emphasize for the heritability estimations: while we found five RBS-R traits to have greater heritability estimates than the autism (PGC) case-control heritability, the confidence intervals around our estimations overlap with the autism (PGC) heritability. Future work with greater sample sizes are necessary for more precise estimates of these common genetic effects, with our power analyses indicating that doubling the our current sample size to be sufficient for 80% power. Second, the DCDQ, RBS-R, SCQ, and CBCL were filled out by the parent or legal guardian on behalf of their child. This leaves potential for several biases, including sex-specific biases (mother vs. father reporting). Third, our analyses only considered common genetic variants as measured by SNPs. However, some *de novo* and/or rare variants (which were not considered in our analyses) can have a strong impact on autism risk [5, 6].

In conclusion, our results show that autistic traits (as characterized in an all-autistic cohort) are genetically heterogeneous: the clinical autism subscale traits are variable in SNP-heritability, PGS associations, and significant PGS by sex interactions. Furthermore, of the three instruments investigated, the RBS-R shows the greatest evidence of genetic signal in both (1) autistic samples (greater heritability) and (2) general population samples (via strongest PGS associations). Although historically the path for increased power GWAS discovery has been a quest for ever-larger case-control sample sizes, our results indicate that investing in richer phenotypic characterization, even at the expense of sample size, might be a better value proposition for psychiatric genetics and autism research in particular.

### 4.1 Language choices

Many autistic self-advocates prefer identity-first language (i.e. autistic individuals), and some autistic individuals and their families prefer person-first language (i.e. individuals with autism). We chose to use identity-first language for this paper. Gender is distinct from sex and has important variance, especially in the autistic community [86, 87]. For language around sex and gender, we exclusively considered sex (meaning sex designated at birth).

## Data Availability

The SPARK data can be obtained at SFARI Base and the ABCD data can be obtained at the ABCD Data Repository. The autism subscale summary statistics, supplementary data, and the code for all analyses can be found at the Michaelson Lab GitHub.

https://base.sfari.org

https://nda.nih.gov/abcd

https://research-git.uiowa.edu/michaelson-lab-public/autism_subscales_common_genetics

## Acknowledgments

We are grateful to all of the individuals and families in SPARK, the SPARK clinical sites, and SPARK staff. We appreciate obtaining access to genetic and phenotypic data for SPARK data on SFARI Base. We are also grateful to all the individuals and families in ABCD.

## Data availability statement

The SPARK data can be obtained at SFARI Base: https://base.sfari.org

The ABCD data can be obtained at the ABCD Data Repository: https://nda.nih.gov/abcd

The autism subscale summary statistics, supplementary data, and the code for all analyses can be found at https://research-git.uiowa.edu/michaelson-lab-public/autism_subscales_common_genetics

## Author contributions

The study was designed by TRT and JJM. The data was preprocessed by TRT, TK, and EB. The analyses were performed by TRT and JJM. The manuscript writing was done by TRT, LGC, AJT, and JJM.

